# A global analysis of patterns of tuberculosis exposure and transmission

**DOI:** 10.1101/2025.10.09.25337649

**Authors:** Peter J. Dodd, Katherine C. Horton

## Abstract

Tuberculosis (TB) is a major public health concern and the leading infectious cause of mortality globally. The disease exhibits strong prevalence patterns by age and sex, but the implications of these patterns for likely TB exposure and transmission have not previously been systematically assessed. We combined estimates of social mixing patterns and TB prevalence for 177 countries to estimate the proportion of TB exposure to and transmission from age groups and sexes. We found that a majority of TB transmission, in both sexes, and for both children and adults, is attributable to contact with adult men. Across age groups, TB exposure typically peaked in adolescence, whereas contributions to TB transmission was flatter or increasing with age, and more variable across regions. Our analysis highlights an important and under-appreciated contribution to transmission in some settings from older adults, who may face particular barriers to healthcare access. More systematic analyses focusing on understanding the epidemiology of TB transmission should be used to inform context-specific prioritization of interventions.

## Introduction

Tuberculosis (TB) remains a major cause of morbidity and mortality worldwide, with an estimated 10.8 million people falling ill and over 1.2 million people dying directly from the disease in 2024 (1). TB affects people of all sexes and age groups, but disease burden is unequally distributed across population groups. National TB prevalence surveys have consistently shown higher disease burden among men relative to women (2). TB prevalence also increased with age in many surveys, peaking in people over age 65 years, though in some settings, prevalence was more evenly distributed across middle and older age groups (1).

While patterns of disease burden identify population groups requiring diagnostic and treatment services, patterns of exposure and transmission are key for targeted prevention strategies to interrupt transmission. Social contact data have helped understand transmission of various respiratory infections in a way that has guided responses, although the focus of most social contact surveys has been on mixing patterns by age, with fewer describing mixing by sex (3). Under the slogan ‘know your epidemic, know your response’ (4), similar analyses have guided the HIV response. This cornerstone approach to policy development has supported HIV programmes to develop targeted, context-specific strategies to prevent new infections and care for those needing treatment (5, 6). For TB, however, there has been very limited systematic and comparative analysis of patterns of exposure and transmission across population groups. These analyses are needed to inform context-specific prioritization of interventions to interrupt transmission.

## Results

By combining estimates of age- and sex-assortative mixing, World Health Organization (WHO) estimates of TB disease burden, and United Nations demographic data, we calculated the relative force-of-infection in and from each sex (female and male) and age group (0-4, 5-14, 15-24, 25-34, 35-44, 45-54, 55-64, and 65+ years) for 2023 in 177 countries, comprising 97.6% of the world’s population and >99.9% of WHO-estimated TB incidence. We aggregated exposures to calculate exposure and transmission by sex and age, globally and by WHO region.

Both exposure and transmission were higher in males than in females. We estimated 61% (95% uncertainty interval [UI]: 58% to 63%) of global transmission was attributable to contact with men aged 15+ years, including 67% (95%UI: 62% to 71%) of transmission to men, 52% (95%UI: 49% to 56%) to women, and 63% (95%UI: 58% to 68%) to children. A majority of global exposure was from contact with men across all age groups and both sexes (Figure 1, panel A). We estimated that if men had the same TB disease levels as women, TB exposure would reduce by 25% (95% UI: 24% to 26%) globally.

**Figure 1.**
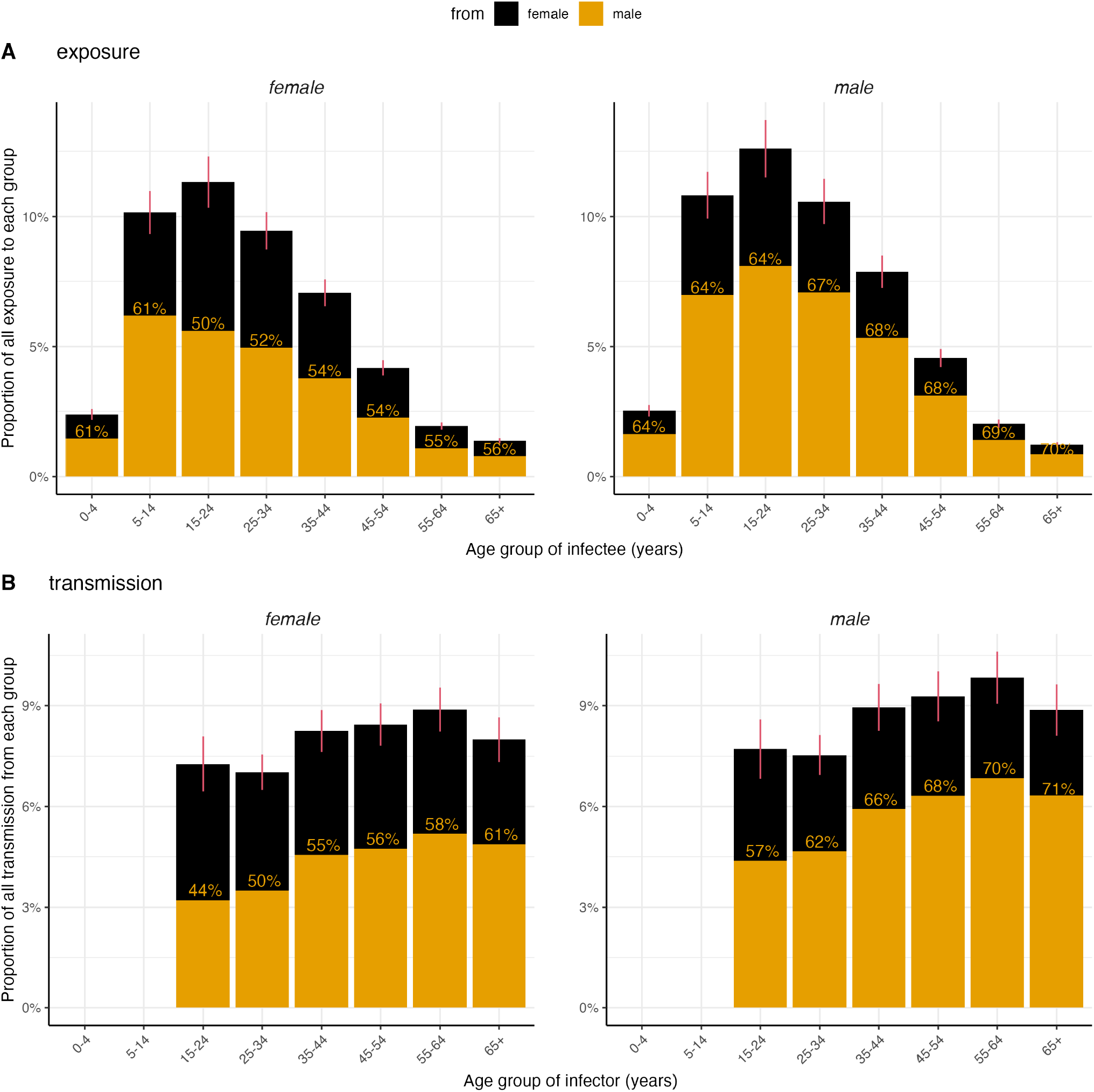
Proportion of global TB exposure (A) and transmission (B) by age and sex in 2023. Left and right columns represent the sex of individuals exposed. Colors represent contributions by sex of those transmitting. Panel A shows the proportion of exposure by age of those exposed. Panel B shows the proportion of exposure by age of those transmitting. Bars sum to 100% across both columns for panel A and panel B separately. Percentage figures show the proportion contribution in each bar from males. Red error bars represent 95% uncertainty intervals.

We found very different patterns of global exposure and transmission by age (see Figure 1). The age of TB exposure peaked around adolescence, before declining into older ages (Figure 1, panel A), whereas contributions to transmission were zero from children and much flatter across adult age groups, highest in the 55-64 year age group (Figure 1, panel B). Globally, 24% (95%UI: 22% to 25%) of TB exposure was in adolescents aged 15-24 years, compared with 7% (95%UI: 6% to 7%) in adults aged 55+ years. By contrast, 15% (95%UI: 14% to 16%) of TB transmission was from adolescents aged 15-24 years, compared with 36% (95%UI: 34% to 37%) from adults aged 55+ years.

While exposure and transmission patterns by sex were consistent across regions, age patterns varied, especially for transmission (Figure 2). The proportion of exposure by age was relatively stable across WHO regions, most commonly peaking in adolescents 15-24 years of age, but peaking in the 5-14 year age group in the Africa region and the 25-34 year age group in the Europe and Western Pacific regions. However, the pattern in contribution to transmission by age varied substantially across regions with the highest proportions due to adolescents 15-24 years of age in the Americas and South-East Asia, the 35-44 year age group in the Africa and Europe regions, and the oldest age groups in the Eastern Mediterranean and Western Pacific regions.

**Figure 2.**
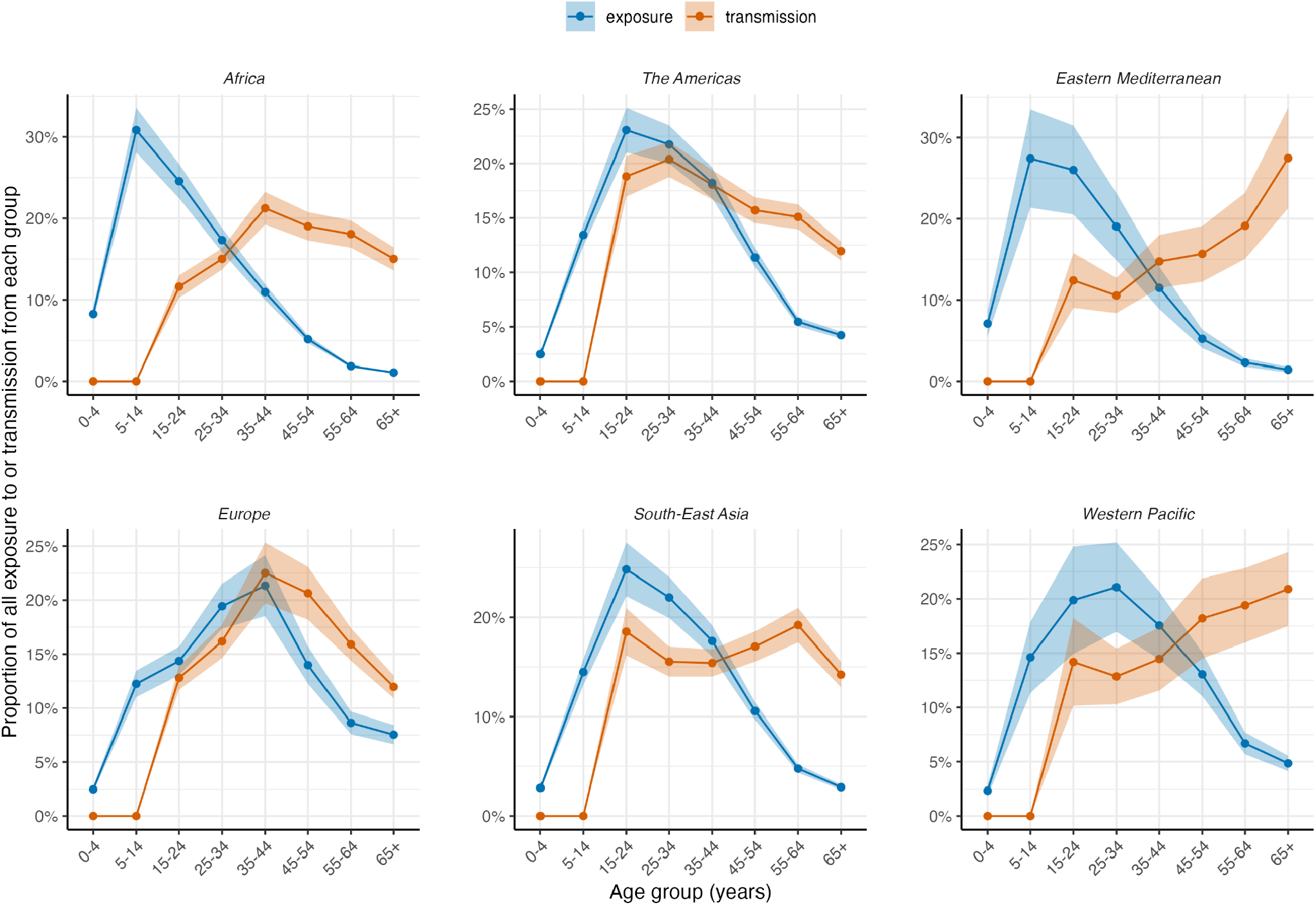
Proportion of all TB exposure (blue) and transmission (red) by age and sex in each WHO region in 2023. Ribbons represent 95% uncertainty intervals.

## Discussion

Our findings highlight heterogeneity in TB transmission and exposure across sexes and age groups. This analysis provides an example of a standardised approach that could be developed to inform priority-setting by providing context-specific information on TB transmission epidemiology.

We found that across all ages globally, and in most settings, men are the majority contributors to TB transmission. This confirms and extends previous work (7). The sex-assortativity in social contact patterns observed across a range of settings has the potential to amplify sex disparities in TB burden driven by underlying differences in biological, sociobehavioural, and health system factors (8). Our results underscore the crucial importance of ensuring TB interventions reach men to reduce transmission to all (9).

Our result that contributions to transmission peak in older ages, especially in the Eastern Mediterranean and Western Pacific regions has, to our knowledge, not been noted previously. This result is driven by patterns in per capita TB prevalence by age (10). Yet in the Americas and South-East Asia, the greatest transmission contribution comes from ages 15-24 years, and in the Africa and Europe regions, from ages 35-44. These findings highlight the need for setting-specific data and analyses to understand exposure and transmission dynamics.

The finding that people aged over 65 years contribute substantially to transmission in some regions has important implications for policy. Demographic change has the potential to slow declines in TB incidence (11) and means that older people will represent an increasing proportion of TB incidence in the future, yet senior age groups may face particular barriers to accessing care (12, 13). More work is required to improve TB services to senior age groups, not just because they are at higher risk of poor outcomes, but because of their contribution to ongoing transmission.

Exposure to TB infection follows a very different pattern, and peaks in adolescence due to increasing contact rates with adult age groups with higher prevalence of infectious TB. This has implications in considering the use of existing and emerging vaccines for TB. If vaccines are administered neonatally, substantial duration of efficacy is required if they are to continue to protect young adults. Routine vaccination of 9-year olds with novel vaccines would provide a boost in immunity as children enter this phase of increased exposure (14).

Our analysis had limitations, including the assumption that adult groups had equal infectiousness and duration of TB (in using incidence as a proxy for prevalence). We considered infection incidence proportional to exposure in the same way across groups, neglecting any differences due to protection against infection conferred by previous TB exposure. Our zero contribution to transmission from children is the result of the assumption that TB in these age groups is not infectious. While this is well-supported for the under 5 year age group, it is likely that there is an increasingly adult pattern of TB disease through the 5-14 year age group, and that the contribution to TB transmission from this group is low but non-zero. Finally, data to inform sex-assortativity and its dependence on age were limited.

Nevertheless, our analysis highlights underappreciated and context-specific patterns in exposure and transmission that are directly relevant to intervention development. Men and older people make substantial contributions to TB transmission in many settings and may require specifically-designed approaches to improve access to care. In most settings adolescents have the highest TB exposure rates, requiring preventive strategies that are able to cover this period. Specific patterns vary by region and country, implying optimal intervention strategies will vary by setting.

## Materials and Methods

We combined updated estimates of age-specific social contact patterns for 177 countries from Prem et al. (15) with a systematic review of sex-assortiativity in social mixing (3). We modelled the contact rate to age group *a* and sex *s* (contactee) from age group *a*′ and sex *s*′ (contactor), *C*_*a,a′*_^*s,s′*^, as

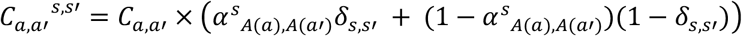

where *δ*^*s,s′*^ is the Kronecker delta that is 1 when *s* = *s*′ and 0 otherwise, and *A*(*a*) maps the finer 5-year age categories (*a*) of Prem et al. (15) onto the 3 age categories in Horton et al. (3) for sex-assortativity.

For country c, the force-of-infection from age group *a*′ and sex *s*′ *to* age group *a* and sex *s* is then

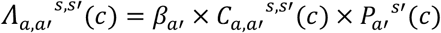

where *P*_*a′*_^*s′*^(c) is the per capita TB prevalence in age group *a*′ and sex *s*′ for country c, and *β* is a transmission probability factor that is assumed constant across all age groups, sexes, and countries, except that it is zero in child age groups (under 15 years of age), which are assumed non-infectious. We use the 2024 WHO estimates of TB incidence in 2023 as a proxy for prevalence, and United Nations World Population Prospect 2024 demographic estimates for 2023. We further aggregated these inputs to a common set of age groups (0-4, 5-14, 15-24, 25-34, 35-44, 45-54, 55-64, and 65+ years) by summing midpoints and variances representing uncertainty.

In calculating aggregate fractions of transmission between sexes and age groups, we average over countries, c, weighting by their populations *N*_*a*_^*s*^(c) in each age group exposed. For example, the fraction of infections from age group *a*′ and sex *s*′ globally *F*_*a′,s′*_ would be given by:

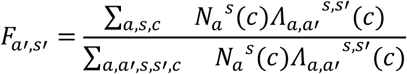

We propagated uncertainty from data inputs and parameters using error propagation. All analyses were performed using R version 4.5.0. Code and data to reproduce these results is available on GitHub at https://github.com/petedodd/ttcontrib.

## Data Availability

All data produced are available online at https://github.com/petedodd/ttcontrib

## Acknowledgments

PJD acknowledges research funding through The UK-South East Asia Vaccine Manufacturing Research Hub. This research is funded by the Department of Health and Social Care using UK Aid funding and is managed by the EPSRC. The views expressed in this publication are those of the authors and not necessarily those of the Department of Health and Social Care. KCH is supported by the UK FCDO (Leaving no-one behind: transforming gendered pathways to health for TB) and the US National Institutes of Health (grant number R-202309-71190). This research has been partially funded by UK aid from the UK government (to KCH); however, the views expressed do not necessarily reflect the UK government’s official policies.

## Author Contributions

Conceptualisation: KCH and PJD. Data collection: KCH and PJD. Formal analysis: PJD. Writing - original draft: KCH and PJD. Writing - review & editing: KCH and PJD.

